# Epidemiology and genetic diversity of SARS-CoV-2 lineages circulating in Africa

**DOI:** 10.1101/2021.05.17.21257341

**Authors:** Olayinka Sunday Okoh, Nicholas Israel Nii-Trebi, Abdulrokeeb Jakkari, Tosin Titus Olaniran, Tosin Yetunde Senbadejo, Anna Aba Kafintu-kwashie, Emmanuel Oluwatobi Dairo, Tajudeen Oladunni Ganiyu, Ifiokakaninyene Ekpo Akaninyene, Louis Odinakaose Ezediuno, Idowu Jesulayomi Adeosun, Michael Asebake Ockiya, Esther Moradeyo Jimah, David J. Spiro, Elijah Kolawole Oladipo, Nídia S. Trovão

## Abstract

COVID-19 disease dynamics have been widely studied in different settings around the globe, but little is known about these patterns in the African continent.

To investigate the epidemiology and genetic diversity of SARS-CoV-2 lineages circulating in Africa, more than 2400 complete genomes from 33 African countries were retrieved from the GISAID database and analyzed. We investigated their diversity using various clade and lineage nomenclature systems, reconstructed their evolutionary divergence and history using maximum likelihood inference methods, and studied the case and death trends in the continent. We also examined potential repeat patterns and motifs across the sequences.

In this study, we show that after almost one year of the COVID-19 pandemic, only 143 out of the 782 Pango lineages found worldwide circulated in Africa, with five different lineages dominating in distinct periods of the pandemic. Analysis of the number of reported deaths in Africa also revealed large heterogeneity across the continent. Phylogenetic analysis revealed that African viruses cluster closely with those from all continents but more notably with viruses from Europe. However, the extent of viral diversity observed among African genomes is closest to that of the Oceania outbreak, most likely due to genomic under-surveillance in Africa. We also identified two motifs that could function as integrin-binding sites and N-glycosylation domains.

These results shed light on the evolutionary dynamics of the circulating viral strains in Africa, elucidate the functions of protein motifs present in the genome sequences, and emphasize the need to expand genomic surveillance efforts in the continent to better understand the molecular, evolutionary, epidemiological, and spatiotemporal dynamics of the COVID-19 pandemic in Africa.

## Introduction

The severe acute respiratory syndrome coronavirus 2 (SARS-CoV-2) that was first reported in Wuhan, China in December 2019, emerged as a novel virus causing a cluster of unusual pneumonia cases. Its outbreak soon became a worldwide pandemic that resulted in a global public health emergency (Guo et al., 2020). Coronavirus disease 2019 (COVID-19) caused by SARS-CoV-2 has affected all seven continents, with Africa currently being the least stricken by the pandemic (Lone and Ahmad, 2020). The first case confirmed in Africa was in Egypt on February 14, 2020, followed by Algeria on February 25, 2020. The first case reported in sub-Saharan Africa was confirmed in Nigeria on February 27, 2020 (NCDC, 2020). The first cases in other African countries were recorded in March 2020 (AfricaCDC, 2020), including in Ghana on March 12, 2020. By May 2, 2021, almost 4.57 million confirmed cases and more than 122 thousand deaths were reported by Africa CDC, as part of the more than 152 million confirmed cases and more than 3.19 million deaths reported globally by WHO.

SARS-CoV-2 is the seventh coronavirus known to infect humans (Corman et al., 2020), and it is the third novel coronavirus known to have caused large-scale outbreaks in the twenty-first century. The first was the SARS-CoV in 2003 that also emerged in China (Alanagreh et al., 2020), followed by the Middle East Respiratory Syndrome Coronavirus (MERS-CoV) (Alanagreh et al., 2020, Cui et al., 2019, Zaki et al., 2012) that emerged in Saudi Arabia in 2012. Unlike SARS-CoV and MERS-CoV that cause severe disease in humans (Kumar et al., 2020), the novel SARS-CoV-2 is more infectious but appears to have a lower case fatality rate (CFR) (Zhang and Holmes, 2020). The genome of SARS-CoV-2 is a positive-sense single-stranded RNA (+ssRNA) of approximately 29.9 kilobases (29891 nucleotides) encoding 9860 amino acids (Sapkota, 2020). The virus is classified among the Betacoronaviruses (β-CoVs) group under the Coronavirinae subfamily of the Coronaviridae family. The β-CoVs genus are known to infect humans, bats, and other wild animals (Chen et al., 2020). Genome replication produces two large ORFs that are translated into polyproteins processed post-translationally to produce 16 proteins comprising four structural proteins, namely envelope (E), spike (S), membrane (M) and nucleocapsid (N), and at least nine accessory proteins, some of which are unique to SARS-CoV-2 and others conserved among coronaviruses (Wu et al., 2020). These 16 proteins play a crucial role in the viral RNA synthesis and immune evasion (Snijder et al., 2020, Shereen et al., 2020). Of the four structural proteins, the S-protein, which is made up of S1 and S2 domains, is known to play a unique role in SARS-CoV-2 replication. The protein functions during host cell attachment and entry by primarily mediating binding to the extracellular domains of its receptor, the angiotensin-converting enzyme 2 (ACE2), a transmembrane protein that is also used by SARS-CoV for cell entry (Wan et al., 2020). SARS-CoV-2 binding to ACE2 and fusion with cellular membrane are facilitated by the S1 receptor-binding domain (RBD) and the S2 subunit, respectively. This unique function makes the spike glycoprotein a target for the development of antibodies, therapeutics, and vaccines. Therefore, the mutational patterns in the S-protein and their circulation trends warrant surveillance for effective interventions as the ongoing COVID-19 pandemic develops.

Protein motifs are small regions of amino acid sequences that facilitate the function of the protein and protein-protein interactions (PPIs). They mediate interactions with cellular proteins and molecular processes within the host cells (Sobhy, 2016). Infection by the virus involves a large number of PPIs between the infective virus and the target host cell (Alguwaizani et al., 2018). Motifs are nucleotide or amino acid sequences that are significant in the genome structure formation, function and conserved regions in protein molecules. The conserved amino acid sequence may be responsible for protein-substrate binding, determines the active domain for enzymatic cleavage, transcription factors and the plasticity of protein, and thus repeated motifs are an essential evolutionary mechanism. Hence, identifying motifs and their repeated patterns is important in determining the binding domain of SARS-CoV-2 and to elucidate the evolutionary relationships among sequences (Luo and Nijveen, 2014). Hence, studying and understanding the functional motifs and repeat patterns of SARS-CoV-2 may aid the prediction of protein characteristics, virus-host protein interactions or other putative roles.

It is noteworthy that RNA viruses, including SARS-CoV-2, commonly generate and accumulate mutations in their genomes. In humans, the high immunological pressure facilitates the accumulation of mutations as the epidemic persists. Mutations in the S protein constitute a major cause of public health concern, as they have the potential to alter the viral tropism and thereby potentially confer adaptation to new hosts, influence transmissibility, clinical outcomes, and/or confer resistance to neutralizing antibodies (Sui et al., 2008, Wibmer et al., 2021). In February 2020, a non-synonymous mutation was detected in the S-protein of SARS-CoV-2 viruses sampled from individuals in China and Europe. The mutation caused an amino acid change from aspartic acid to glycine at position 614 (D614G). Experimental and clinical findings have associated the G614 variant with a selective advantage over the D614 virus, resulting in higher viral loads and increased infectivity (Korber et al., 2020), but not necessarily increasing the mortality rate (Plante et al., 2021). In late Summer to early Autumn of 2020, several new variants of the SARS-CoV-2 virus emerged. Variant 20B/501Y.V1 202012/01 classified as Pango lineage B.1.1.7, was identified in the United Kingdom (UK) and designated a Variant of Concern (VOC) (Volz et al., 2021). This variant emerged with an unusually large number of mutations and has since been detected in numerous countries around the globe, including several in Africa. Almost simultaneously, the independent emergence of the VOC 20C/501Y.V2 belonging to Pango lineage B.1.351 was detected in South Africa (Tegally et al., 2021). Cases attributed to this variant have since been detected outside of South Africa (Volz et al., 2021), including disseminating northwards in Africa. Early in 2021, the VOC 20J/501Y.V3 classified as Pango lineage P.1 was first identified in Brazil (Faria et al., 2021)and rapidly spread throughout the Americas, Europe, and Oceania. Despite the independent emergence of the 20B/501Y.V1, 20C/501Y.V2 and 20J/501Y.V3, they share a few common mutations. Since most of the current SARS-CoV-2 immunotherapeutic strategies target blocking the RBD of the S-protein to prevent the binding of SARS-CoV-2 with ACE2 (Chan et al., 2020), alterations in the S-protein sequence could potentially affect the efficacy of immune-based therapeutic agents (Wibmer et al., 2021), making surveillance of spike mutations imperative to aid in the development of effective pharmaceutical interventions.

As of January 7, 2021, nearly one year since the first case was reported in Africa, a total of 5229 SARS-CoV-2 complete genome sequences from 33 African countries had been deposited in public sequence databases including GISAID, which can be studied to better understand the ongoing molecular epidemiology of SARS-CoV-2 in the continent (Oladipo et al., 2020). Initial genome sequence analysis suggested the importation of multiple SARS-CoV-2 strains, mainly of European origin and partly from China (Tessema et al., 2020).

Knowledge of the evolutionary dynamics underpinning the viral genome allows tracing the ongoing outbreak and informs the development and deployment of diagnostic tests (Wang et al., 2020)as well as of effective antiviral and vaccination strategies (Avise, 2000). For example, a recent genome-wide association study on SARS-CoV-2 genomes found variations at the genomic position 11083 within the coding region of non-structural protein 6 to be associated with COVID-19 severity. The study showed that the G11083 variant was more commonly found in symptomatic cases, while the T11083 variant appeared to be more frequently associated with asymptomatic infections (Aiewsakun et al., 2020). Toyoshima and colleagues (2020) also performed a comprehensive investigation of 12343 SARS-CoV-2 genome sequences isolated from individuals in six geographic areas and found that ORF1ab L4715 and S protein G614 variants showed significant positive correlations with fatality rates, which supports the finding suggesting that SARS-CoV-2 mutations might affect the susceptibility to SARS-CoV-2 infection or severity of COVID-19 (Toyoshima et al., 2020). It is to be noted that most sequences from Africa included in the genome variation analysis described above were mostly from North African countries including Egypt, but not those of sub-Saharan Africa.

The emerging trend of SARS-CoV-2 in the African continent calls for a comprehensive study of the genomic and evolutionary patterns of this virus. Comparative analysis of viral genome sequences represents a very useful approach to provide insight into pathogen emergence and evolution. This study therefore pursues an in-depth investigation into the epidemiology, evolution, and molecular motifs of SARS-CoV-2 in Africa to shed light on the pandemic dynamics, and aid inform the development and implementation of control measures in the African continent.

## Materials & Methods

### Assembly of study dataset

SARS-CoV-2 genome sequences collected from Africa were obtained from the Global Initiative on Sharing All Influenza Data (GISAID) database (https://www.gisaid.org/) on January 7, 2020, by only selecting complete genomes excluding those with low coverage. As of January 7, 2020, nearly eleven months since the first case was reported in Africa, a total of 5229 SARS-CoV-2 complete genome sequences from 33 African countries were available in the GISAID database. The sequences were aligned using the online version of the MAFFT (Katoh et al., 2019) multiple sequence alignment tool hosted at https://mafft.cbrc.jp/alignment/software/closelyrelatedviralgenomes.html, with the Wuhan-Hu-1 (www.ncbi.nlm.nih.gov/nuccore/MN908947.3) as the reference sequence. The aligned sequences were manually edited and cleaned in AliView version 1.26 (Larsson, 2014). Sequences with fewer than 75% unambiguous bases were excluded, as were duplicate sequences defined as having identical nucleotide composition and having been collected on the same date and in the same country. The resulting dataset was trimmed at the 5’ and 3’ ends. This dataset was subjected to multiple iterations of phylogeny reconstruction using IQ-TREE multicore software version v1.6.12 (Nguyen et al., 2015) with parameters -m GTR+G -nt 50. TempEst (Rambaut et al., 2016) was used to exclude outlier sequences whose genetic divergence and sampling date were incongruent, resulting in a dataset with 2422 sequences with 29796 nucleotide base pairs.

### Selecting a genomic background dataset

We used the Pango lineage classification available in the metadata associated with the sequences to identify the lineages circulating in Africa (B.1.5, B.1, B.1.1, B.1.1.206, B.1.351, C.1, B.1.5.12, A, B.1.1.273, H.1, B.1.237, B.1.1.52, B.1.1.293, B.1.106, and B.1.1.1), as this nomenclature system is designed to integrate both genetic and geographical information about SARS-CoV-2 dynamics (O’Toole et al., in prep). In total, there were 143 Pango lineages circulating in Africa, as listed in Supplementary Table 1. On January 7, 2021, we obtained from GISAID all available sequences belonging to lineages that circulate in Africa. A similar approach to that described above (including alignment using MAFFT, manual inspection using AliView, phylogenetic tree reconstruction with IQ-TREE and exclusion of root-to-tip outliers using TempEst) was employed, resulting in a dataset with 5002 sequences with 29796 nucleotide base pairs.

### Phylogenetic inference

We merged the study and background datasets resulting in a final dataset with 7416 sequences (Supplementary Table 2). We computed the phylogeny with ultrafast bootstraps using IQ-TREE v1.6.12 (Hoang et al., 2018, Nguyen et al., 2015) with parameters -m GTR+G -bb 1000 -bnni –nt 50. These analyses were conducted using the high-performance computational capabilities of the Biowulf Linux cluster at the National Institutes of Health (Bethesda, MD, USA) (http://biowulf.nih.gov). Trees were rooted in the Wuhan-Hu-1 (accession number: MN908947.3) reference genome and visualized in FigTree version 1.4.4.

### Comparison of evolutionary divergence

We estimated the evolutionary divergence of several sequence datasets from each continent. Each continent-specific dataset consisted of sequences from the final dataset described above. The final continent-specific datasets were as follows: Africa (n = 2414); Asia (n = 1008); Europe (n = 999); North America (n = 997); Oceania (n =1000); South America (n = 998).

To estimate the evolutionary divergence, we calculated the pairwise distance (in base substitutions per site) between all pairs of sequences within and between each continent. We conducted the analyses using the Molecular Evolutionary Genetics Analysis software version 10 (MEGA X) (Kumar et al., 2018, Stecher et al., 2020) and applied the maximum composite likelihood mode (Tamura et al., 2004). The rate variation among sites was modeled with a gamma distribution (shape parameter = 4), and the differences in the composition bias among sequences were considered in evolutionary comparisons (Tamura and Kumar, 2002). We included 1st+2nd+3rd+non-coding codon positions, and all with less than 50% site coverage due to alignment gaps, missing data, and ambiguous bases, were eliminated (partial deletion option). R 4.0.3 software (RCoreTeam, 2019) was used for the visualization.

### Geographical distribution of COVID-19 pandemic in Africa

SARS-CoV-2 genetic data was collected from GISAID as described above and epidemiological data was obtained from OurWorldInData.org (Roser et al., 2020) on January 8, 2021. One sequence did not have a clade assignment and was excluded from the statistics. R 4.0.3 software (RCoreTeam, 2019) was used for the statistical analysis. The R packages used were maptools (Bivand et al., 2021), RColorBrewer (Neuwirth, 2014), maps (Becker et al., 2018), mapdata (Becker et al., 2018), readxl (Wickham and Bryan, 2019), ggplot2 (Kassambara, 2019), qwraps2 (DeWitt, 2021), dplyr (Wickham and Wickham, 2020), gridExtra (Auguie, 2017), ggcorrplot (Kassambara, 2019), ggpubr (Kassambara, 2020), gridExtra (Auguie, 2017), tidyr (Wickham and Henry, 2020), scatterpie (Yu, 2018), ggmap (Kahle and Wickham, 2013), and mapproj (McIlroy et al., 2020). The data and the R script for the analysis can be accessed at https://github.com/Yinkaokoh/updatedSARCoV2_project.

### Detection of repeat patterns and motifs

The retrieved SARS-CoV-2 sequences from Africa were annotated using GLAM2 (http://meme-suite.org/tools/glam2). GLAM2 is a deletion and motif finding software for either nucleotide or amino acid sequences (Frith et al., 2008). The Wuhan isolate with accession number NC_045512.2 was annotated for novel motifs, and the Biostrings R package from Bioconductor was used to find the motifs’ appearance in the retrieved African SARS-CoV-2 sequences. The Tomtom tool (http://meme-suite.org/tools/tomtom), which equates one or more motifs against a database of known motifs, was employed to find overlapping positions across the motif database (Gupta et al., 2007).

## Results

### Epidemiological trends of SARS-CoV-2 in Africa

Worldwide, we observed 782 Pango lineages, 9 GISAID clades and 10 Nextstrain clades. Europe submitted about 65% (n = 208,538) of the SARS-CoV-2 sequences in GISAID as shown in Figure 1, the majority being from the United Kingdom (46%, n = 147,137). Africa submitted just 2% of the SARS-CoV-2 sequences (n = 5,229) with most of the sequences (55%, n = 2882) coming from South Africa. Democratic Republic of the Congo and Gambia followed with 7% (n = 360) each, then Kenya with 290 sequences (6%) and Nigeria with 4% (n = 223) (Supplementary Figure 1). Even though in absolute terms South Africa submitted most of the sequences (Figure 1 - left), a closer look at the number of sequences submitted (Figure 1 - right) revealed that the Democratic Republic of the Congo was the largest contributor of genomic data per 1,000 reported COVID-19 cases in African countries.

**Figure 1:**
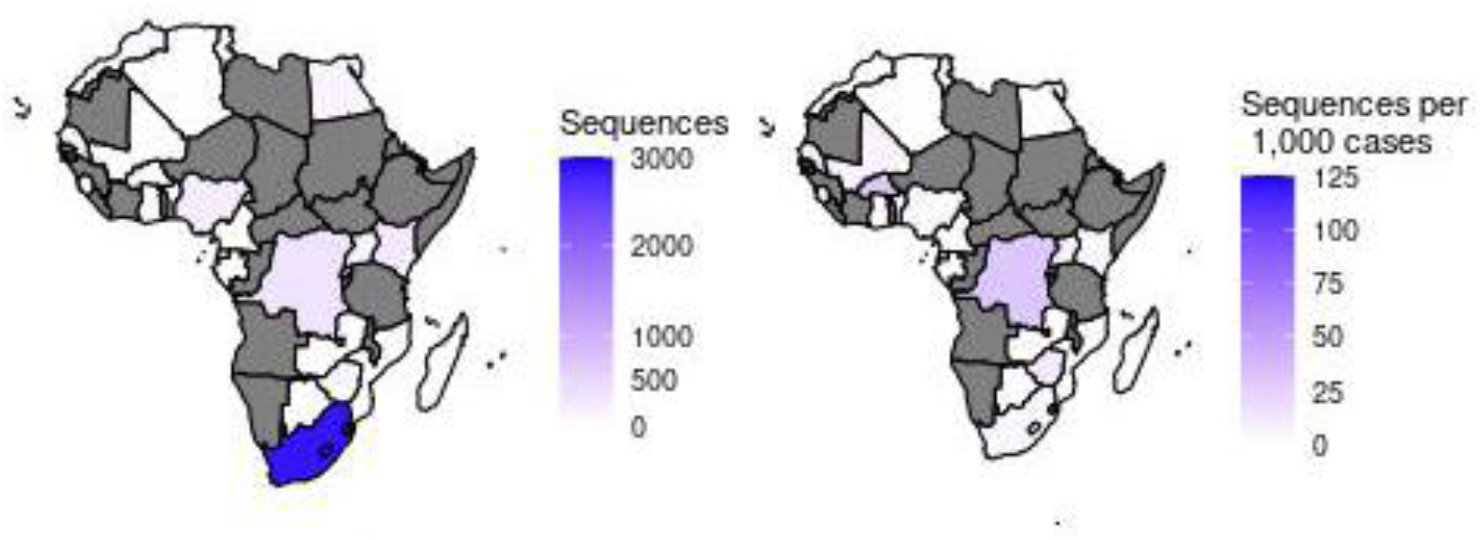
Sequences from African countries submitted to GISAID. Absolute number of sequences from Africa submitted to GISAID (left). Sequences available in GISAID per 1,000 SARS-CoV-2 cases in Africa (right). Grey shade represents countries for which no sequences were available (n = 27/57 (47.4%) countries).

Using Nextstrain clade nomenclature (Supplementary Figure 1), we observed that 20A.EU1 is the dominant circulating clade in Europe followed by 20B and 20A. Clades 20C and 20A are predominant in North America, while 20B is predominant in Oceania. In Africa (Supplementary Table 3), 20B and 20A are the dominant circulating clades, accounting for about 82% of all sequences available for the continent.

For simplicity, we present the most prevalent (top 1%) of the 782 Pango lineages identified worldwide in Supplementary Figure 2. Europe has the most diverse lineages with B.1.177 being the most prevalent, followed by B.1.1 and B.1. In Asia, B.1.1 and B.1.1.284 are the dominant lineages, while D.2 is the most prevalent in Oceania. B.1, B.1.2 and B.1.1 are the most prevalent in North America.

Interestingly, the top 1% Pango lineages (Supplementary Figure 2) circulating elsewhere in the world were not observed in sequences from Africa and South America. The top 10% of the lineages circulating in Africa is represented in Supplementary Figure 4. Pango lineage B.1.5 is the dominant lineage circulating in Africa representing 11.3% (n = 591) of all diversity. Other prevalent lineages in Africa are B.1 (n = 546, 10.4%), B.1.1 (n = 518, 9.91%), B.1.1.206 (n = 481, 9.20%), B.1.351 (n = 349, 7%) and C.1 (n = 271, 5%).

The first sequence from Africa was collected on March 1, 2020 and belongs to Pango lineage B.1.5 (Figure 2), while the second SARS-CoV-2 case in Africa, reported on March 8, 2020, belongs to the B.1 lineage. Pango lineages B.1, B.1.1 and B.1.5 circulated in Africa from March 1, 2020 through June 7, 2020. Pango lineage B.1.1.206 was first reported in Africa on June 8, 2020. While B.1.5 was the most prevalent lineage in Africa between March 1 and June 7, it was replaced by B.1.1.206 which was dominant between June 8 and November 2, 2020. The Pango lineage B.1.351 was first reported in South Africa (on October 10, 2020 and it became the most prevalent lineage on November 3, 2020, likely as a consequence of the increased sequencing efforts in the country.

**Figure 2:**
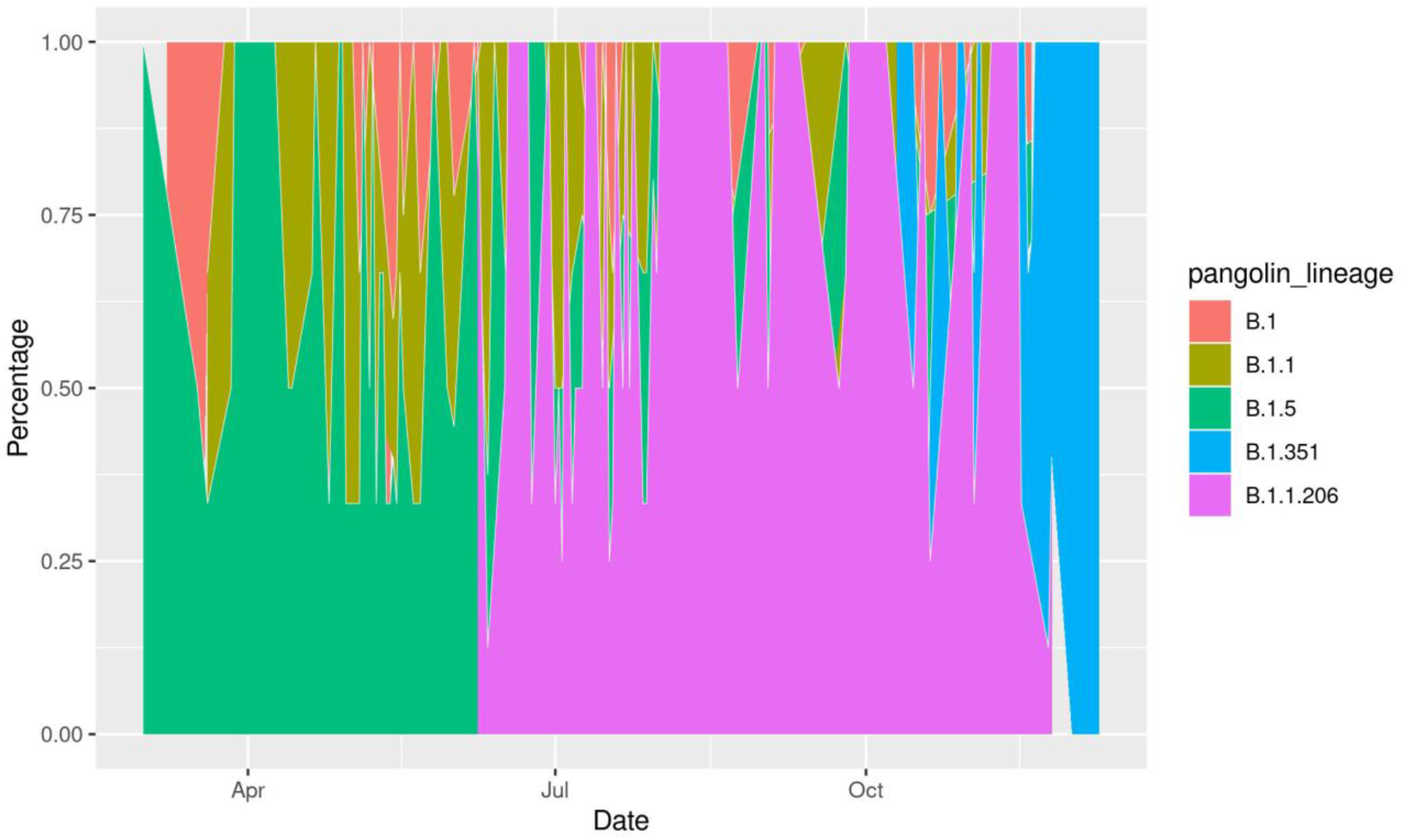
The incidence of the top 5 Pango lineages circulating in Africa between March 1, 2020 and January 7, 2021.

Focusing on data found in GISAID will not give a true representation of the disease dynamics of SARS-CoV-2 in Africa, as only a fraction of the cases in Africa are sequenced. Therefore, we analyzed reported COVID-19 cases using data from OurWorldInData.org downloaded on January 8, 2021. As presented in Supplementary Figure 3, North America has the highest number of COVID-19 cases (n = 25 million), followed by Europe (n = 19 million) and Asia (n = 18 million). Oceania has the least number of cases reported of the six continents (n = 20,575). The absolute number of cases in Africa (n = 2 million) and South America (n = 6 million) are a very small fraction of the cases in Asia, North America, and Europe. The global average of COVID-19 cases per 100,000 population (hereafter referred to as cases/pop) is 895, represented by the red line in Figure 3. We observed that the average number of COVID-19 cases per 100,000 persons in Oceania, Africa, and Asia are all below the global average. Considering the absolute cases of COVID-19, Asia is more affected than South America, however, taking population into consideration reveals that South America (1287 cases/pop) has a higher burden of COVID-19 cases than Asia (390 cases/pop).

**Figure 3:**
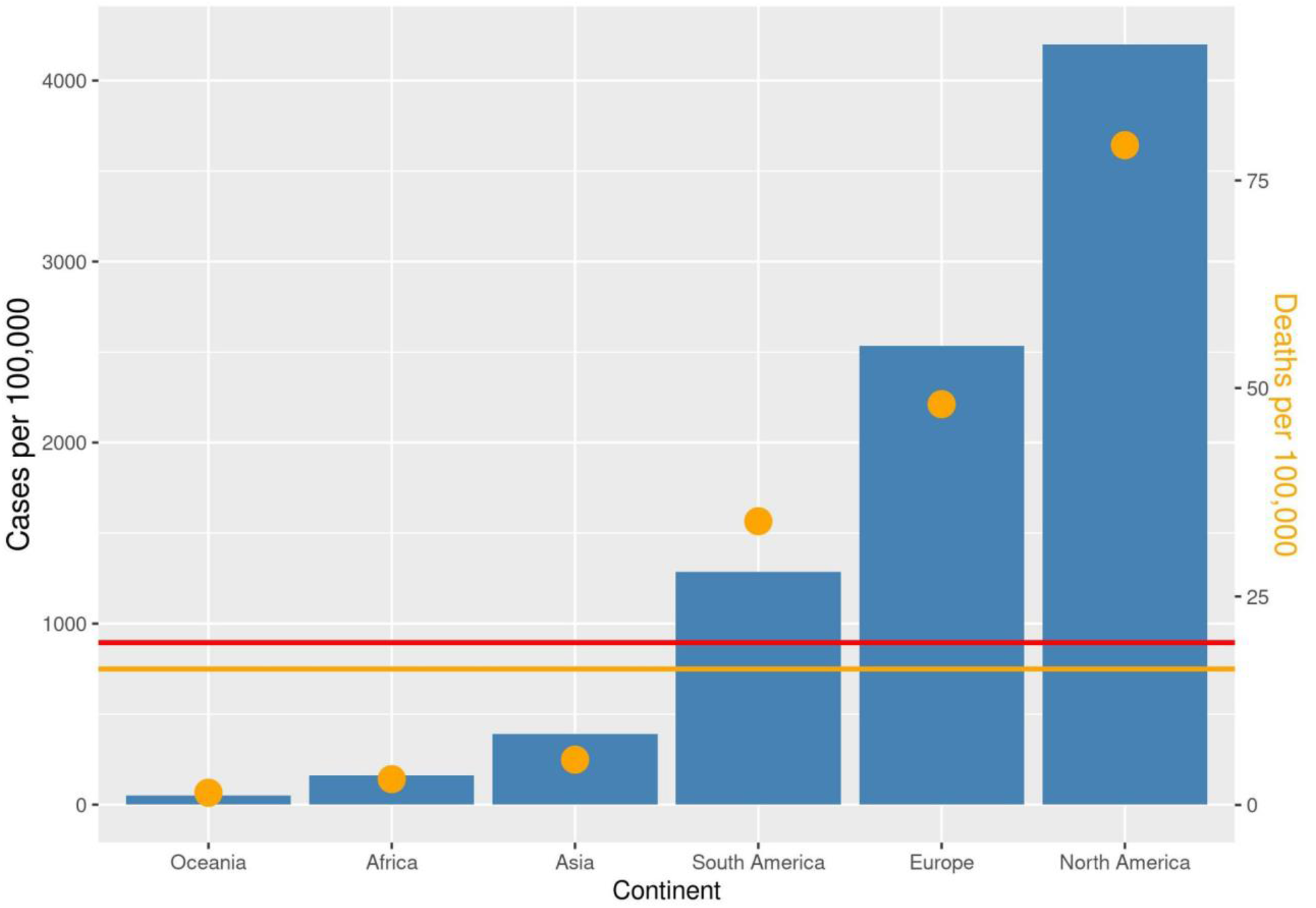
Number of COVID-19 reported cases and deaths per 100,000 population in the different continents. The red line represents the average absolute number of COVID-19 cases worldwide per 100,000 population in the world (i.e (global COVID-19 cases/world population) x 100,000). The orange points represent the average number of deaths per 100,000 population in the different continents with its scale on the left y-axis. The orange line represents the number of deaths globally per 100,000 world population.

The number of deaths per 100,000 population (hereafter referred to as deaths/pop) followed the same trend as the number of cases per 100,000. Deaths per 100,000 in Oceania (2 deaths/pop), Africa (4 deaths/pop), Asia (6 deaths/pop) are far below the global value (19 deaths/pop), while it is above in South America (39 deaths/pop), Europe (55 deaths/pop), and North America (91 deaths/pop). This shows a positive correlation between the number of COVID-19 tests and the number of COVID-19 cases and deaths reported.

In Africa, an analysis of COVID-19 cases per 100,000 population (Figure 4) showed that, while the continent had comparatively low case numbers, individual nations had high COVID-19 burdens South Africa has been the most seriously affected with 1260 cases/pop; however, Tunisia (678 cases/pop) and Morocco (791 cases/pop) in North Africa appeared to also be greatly impacted. Similar patterns were observed for Libya in North Africa (1076 cases/pop), Namibia (528 cases/pop) and Botswana (347 cases/pop) in Southern Africa, and Gabon in Central Africa (404 cases/pop). Other African countries were mildly impacted. Overall, the number of cases/pop demonstrate that northern and southern countries, and Gabon in Central Africa, were the most affected regions/country in Africa.

**Figure 4:**
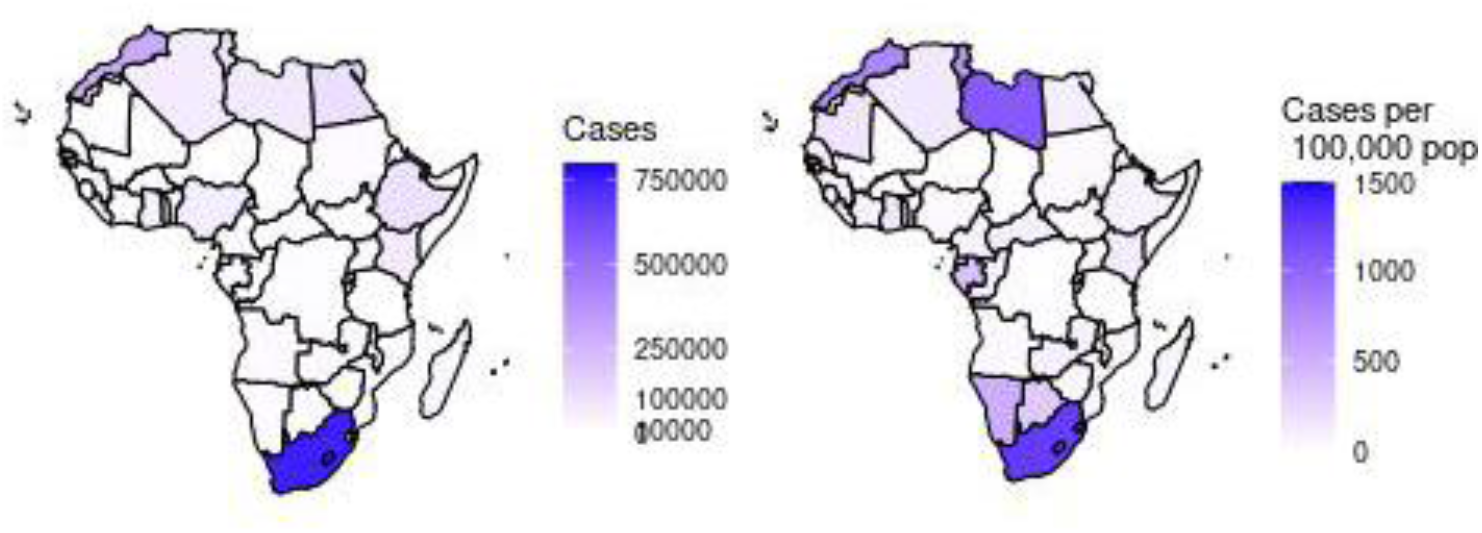
COVID-19 cases reported in African countries. Absolute number of cases (left). Number of cases per 100,000 population (right).

To get a deeper insight into the impact of SARS-CoV-2 on African countries, we analyzed the reported deaths as represented in Figure 5. Considering the absolute number of deaths, South Africa remained the worst hit on the continent with 20241 deaths, distantly followed by Egypt (n = 6453), and other North African countries. While Ethiopia, Kenya, Nigeria, Sudan, and Libya have reported more than 1000 deaths each, about 20 African countries have recorded less than a total of 500 deaths. Considering the number of deaths recorded per 1,000 reported cases (deaths/cases) (Figure 5 - right), we observed that Western Sahara was the worst affected with 100 death/cases, followed by Sudan (76 death/cases), Egypt in the North (58 death/cases) and Chad in the West (63 death/cases).

**Figure 5:**
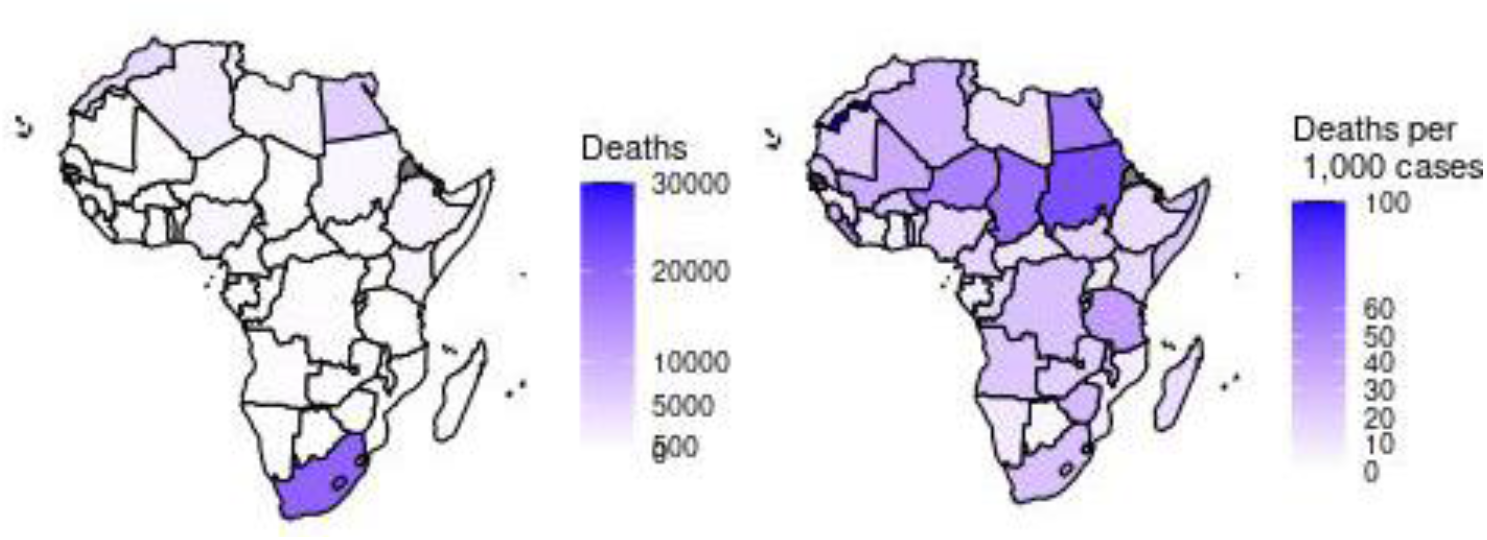
Reported deaths from COVID-19 in Africa. Absolute number of total deaths (left) per country. Number of deaths per 1,000 reported cases (right).

South Africa and Morocco recorded the highest absolute numbers of COVID-19 cases in Africa (Figure 4 - left), though this might reflect the large number of tests conducted in these two countries (Figure 6), 5,110,384 and 3,646,330 for South Africa and Morocco, respectively. The third highest number of tests was conducted in Ethiopia (n = 1,562,008) albeit approximately 60% less than the number of tests conducted in Morocco. Taking population into consideration (number of tests conducted per 100,000 population; tests/pop), the islands of Mauritius and Cape Vert performed the highest number of tests (22,389 tests/pop and 18,250 tests/pop, respectively) followed by Botswana (13,955 tests/pop) and Gabon (11,860 tests/pop) who carried out more tests, relative to population, than South Africa (8,576 tests/pop) and Morocco (9,835 tests/pop).

**Figure 6:**
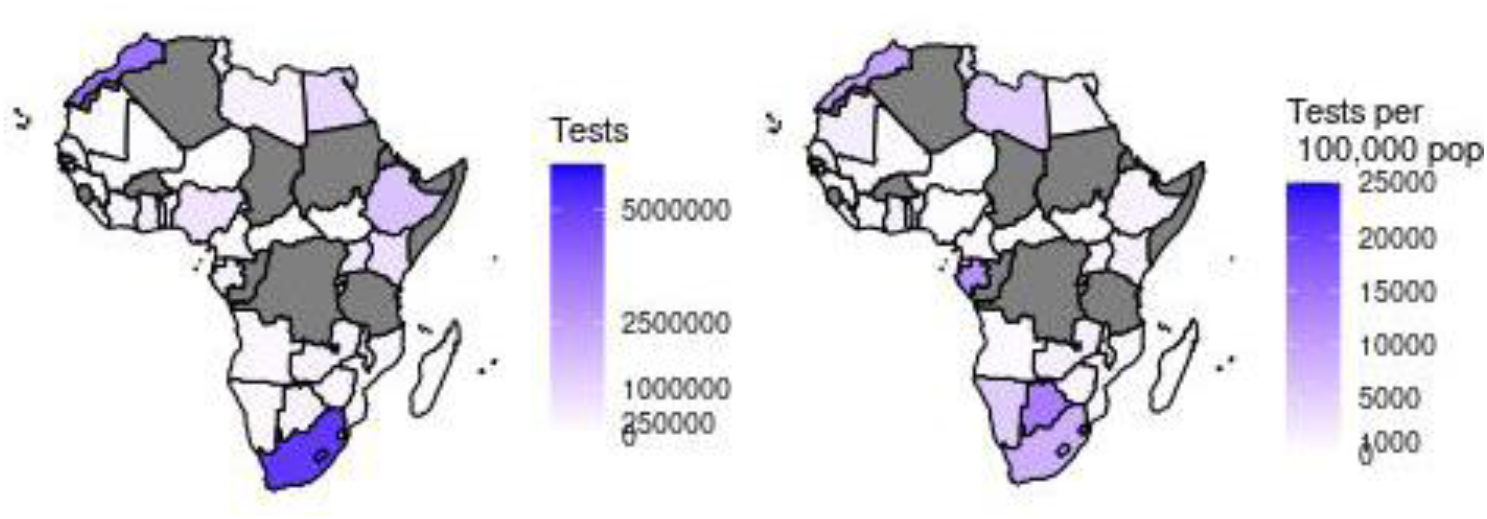
Number of SARS-CoV-2 tests conducted in African countries. Absolute number of tests (left). Number of tests per 100,000 population (right).

We analyzed the number of positive tests per 1,000 COVID-19 tests (pos/test) conducted (Figure 7). Mayotte was estimated to have the highest positivity rate (271 pos/test), followed by Guinea (250 pos/test), South Sudan (249 pos/test) and Tunisia (200 pos/test). Some other African countries such as Libya (198 pos/test), Madagascar (186 pos/test), Gambia (166 pos/test), Cameroon (152 pos/test), Central African Republic (150 pos/test), South Africa (147 pos/test), Reunion (141 pos/test), Sao Tome and Principe (136 pos/test), Swaziland (110 pos/test), Egypt (110) and Ivory Coast (102 pos/test) have more than 100 positive tests per 1,000 COVID-19 tests. The lowest positivity rates were observed in countries such as Benin (10 pos/test), Rwanda (9 pos/test), and Mauritius (2 pos/test).

**Figure 7:**
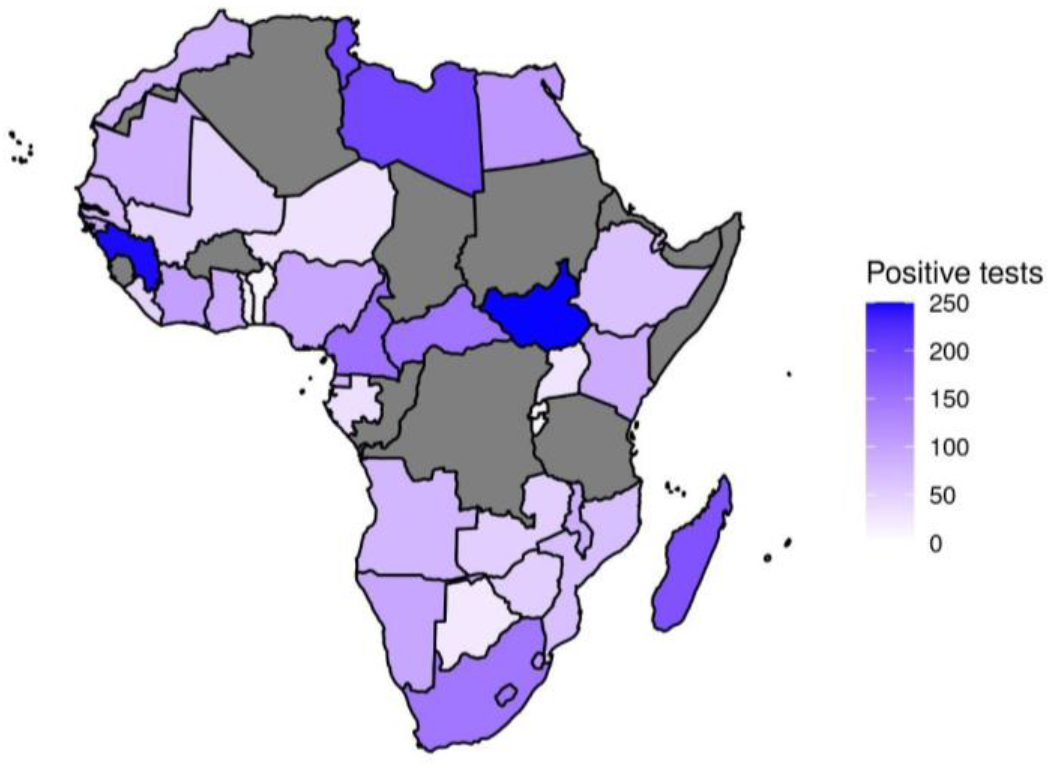
COVID-19 positivity rate in Africa. Number of positive patients out of every 1000 COVID-19 tests. Grey shades represent countries for which data is not available.

### Identification of novel repeat patterns and motifs

GLAM2 (https://meme-suite.org/meme/tools/glam2) determines novel recurring motifs with deletions and insertions, and their presence in functional proteins. Using the Tomtom motif tool (http://meme-suite.org/tools/tomtom), we identified the motifs represented in the table below, as well as their functional class when compared with the motif database (http://elm.eu.org/elms/LIG_IBS_1.html). The p-value denotes the likelihood of a random motif with equal width to function as a target and align better to motifs in the database, thus producing a match score as good or better than another target. The e-value shows the expected number of false positives in the matches, with a threshold of ten or less.

The motifs were matched with the study dataset but were not found in all isolates (data not shown). We observed a position shift in the motifs across the isolates, however it was not clear if the shifts altered the functionality of the motifs. It therefore indicates that the motifs are present but at different positions in the genome, probably due to deletions and substitutions along the genome of African SARS-CoV-2 isolates, as is true for RNA viruses.

Two of the discovered motifs were found in the ORF1ab gene (locus Gu280-gp01) and were identified as integrin binding sites occurring mainly at positions 396-445 (ID 1) and 3361-3409 (ID 2). We also observed a motif that could function as an N-glycosylation site, mostly in positions 396-445 (ID 1).

**Table 1:**
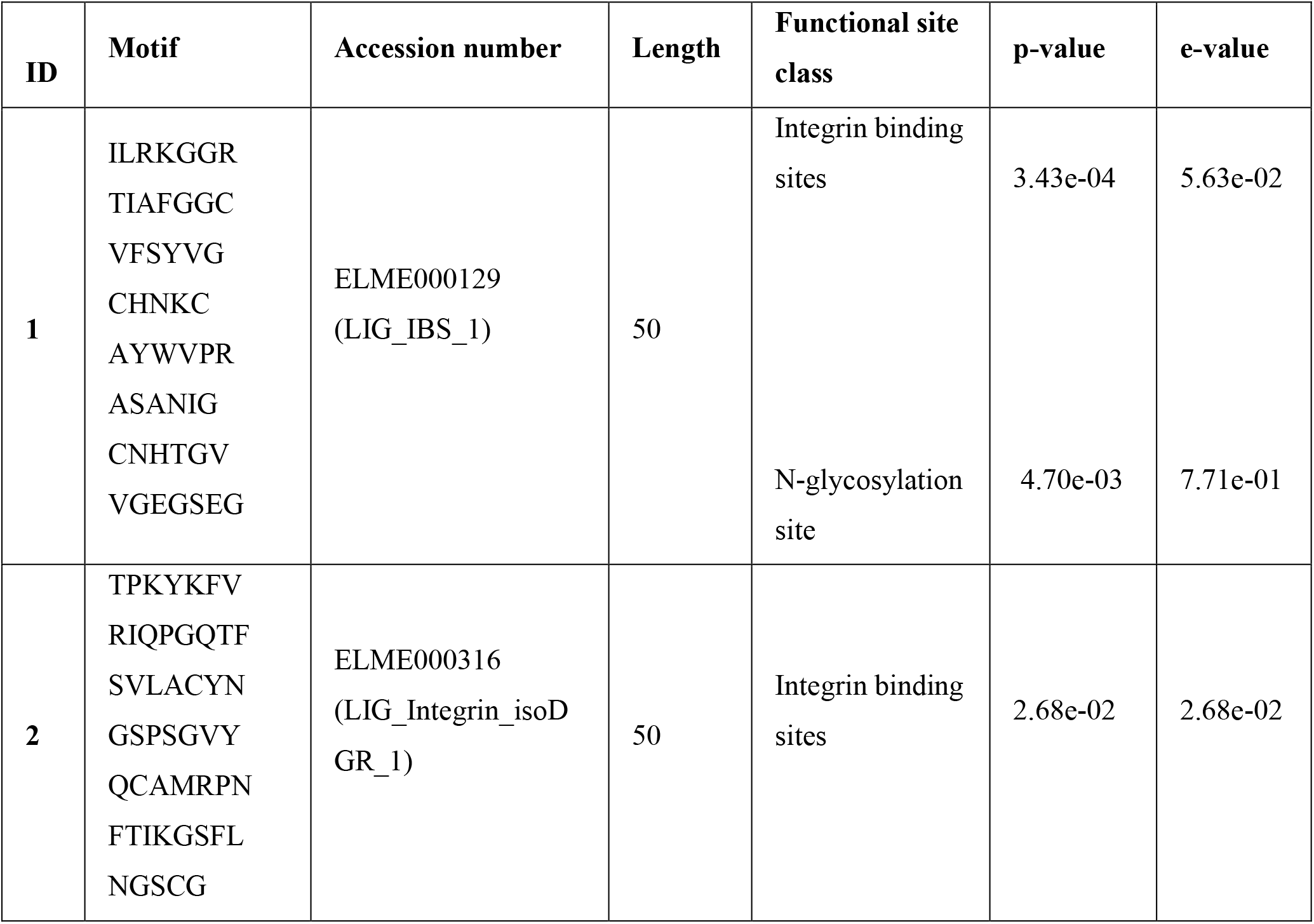
Repeat patterns and motifs in SARS-CoV-2 genomes from Africa.

### Evolutionary history

We analyzed the phylogenetic relationships among African sequences and those from other parts of the world (Figure 8). The phylogenetic tree demonstrates that African viruses cluster closely with viruses from all continents, but mostly with those from Europe, a source that generated some of the large outbreaks detected in the phylogenetic tree. We also observed that African clusters tend to contain mostly sequences from the same or adjacent countries (Supplementary Figure 6). Even though the South African outbreaks have been studied in depth (Giandhari et al., 2021, Tegally et al., 2021, Tegally H, 2021), we also identified outbreaks (clusters with more than 15 sequences) in Egypt, Democratic Republic of the Congo, and Gambia.

**Figure 8.**
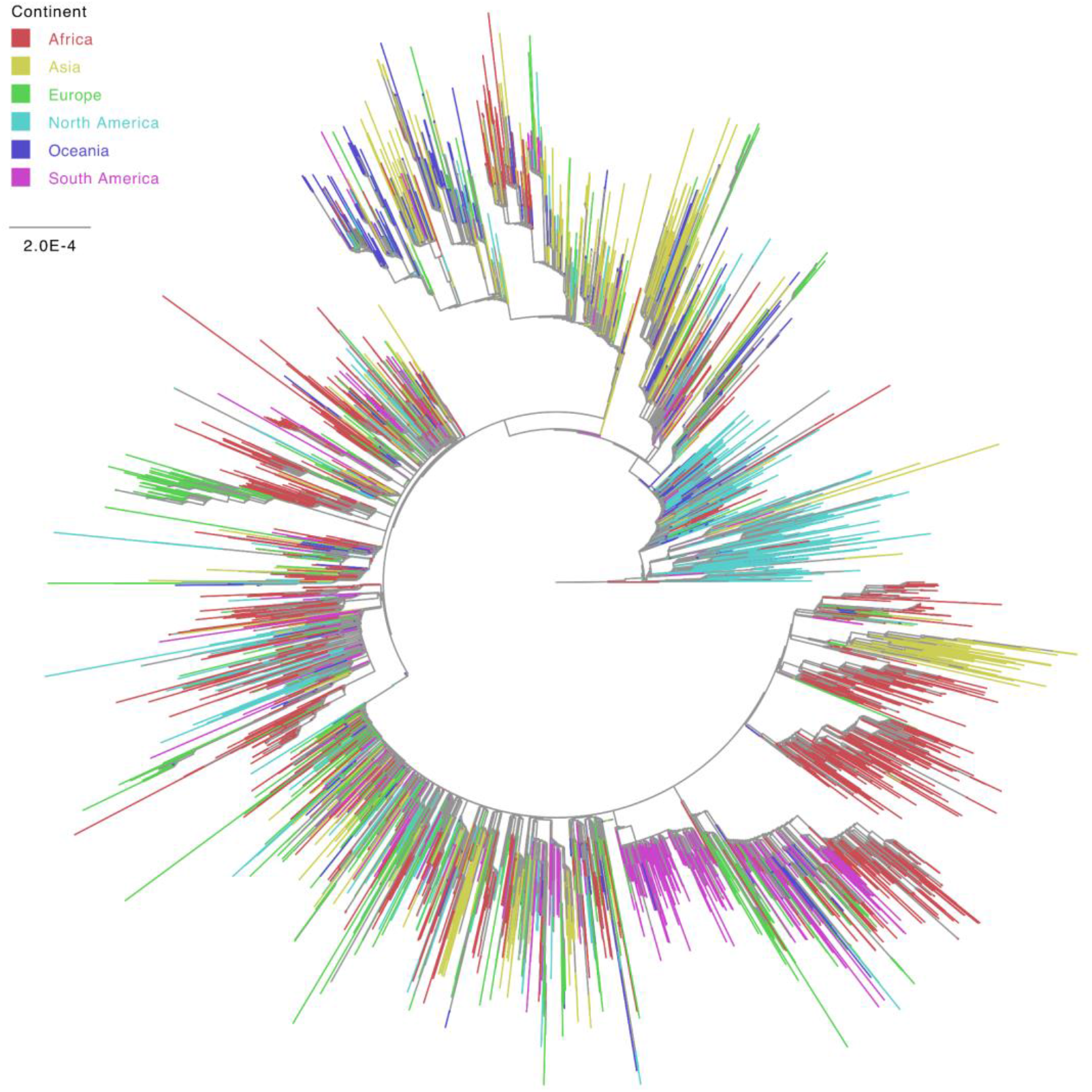
Maximum likelihood tree colored by continent. Phylogenetic tree inferred for a dataset with genetic sequences from all continents.

We also looked specifically at the viral genetic diversity within Africa, as compared to the genetic diversity observed in other continents (Figure 9 and Supplementary Table 4). Inspection of the continent-specific genetic distance distributions with a Wilcoxon signed-rank test revealed that the viral diversity circulating in Africa is significantly higher (p-value < 2.2e-16) than that estimated in Oceania and South America, but significantly lower than that in Asia, Europe, and North America. In line with this, we observed that the African epidemic is closest to that of South America and farthest from those of Asia and North America (Supplementary Figure 7).

**Figure 9.**
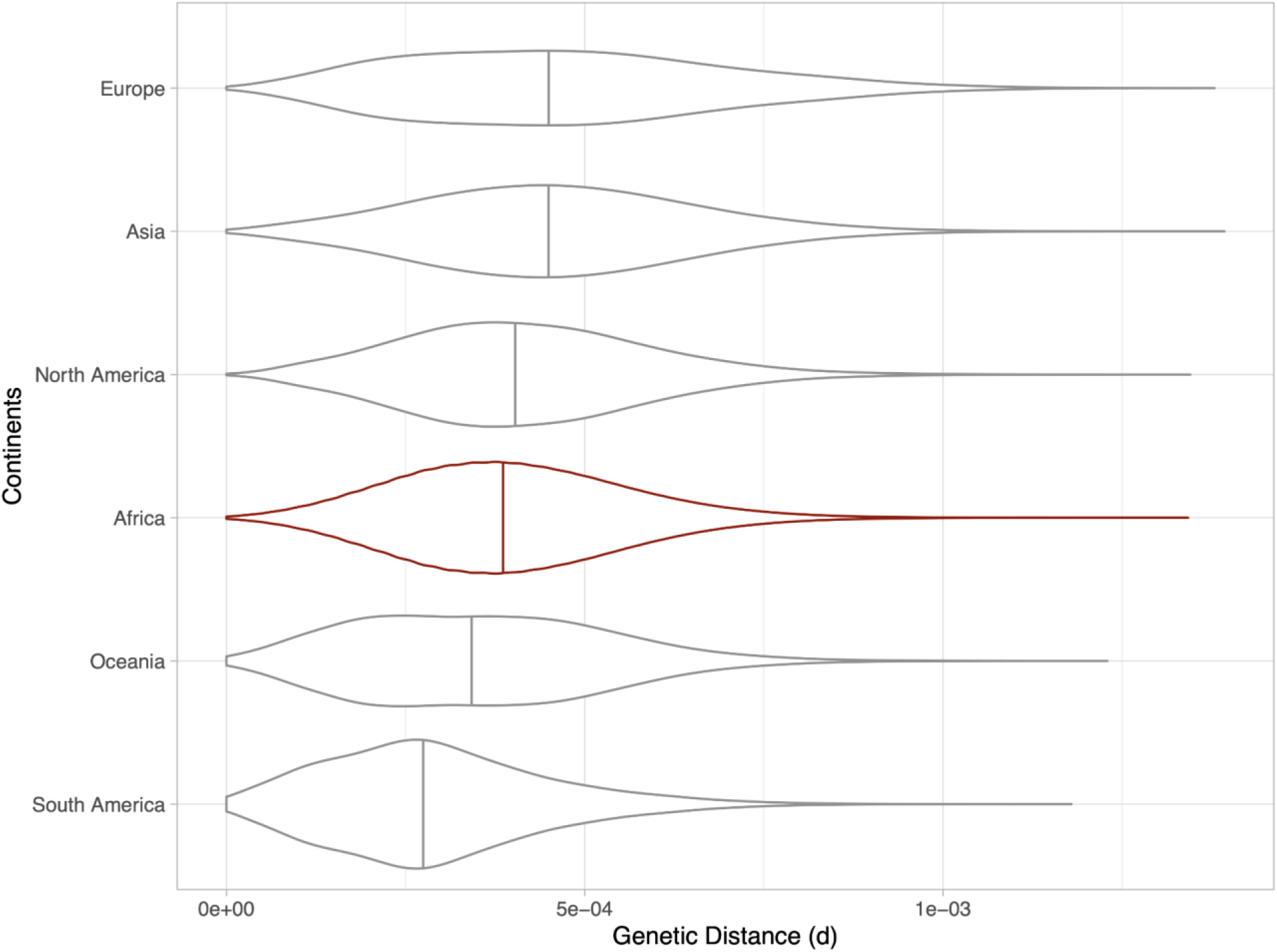
Evolutionary divergence of SARS-CoV-2 across continents. Violin plots represent the distribution of pairwise genetic distances between all sequences for isolates in each continent. Vertical lines depict the mean pairwise genetic distance between all samples in each continent.

## Discussion and Conclusion

The investigation of SARS-CoV-2 classical and genomic epidemiology in Africa is crucial for monitoring the circulating genetic diversity of the virus, its clinical presentation and epidemiological profiles, estimate the magnitude of a pandemic, and to inform the effective development and implementation of control measures in the African continent. Mutations in the viral genome may also raise concerns for reliable and effective detection, and monitoring of the transmission dynamics (Galloway et al., 2021).

Whereas SARS-CoV-2 sequence data constitute an integral part of the decision making in other continents (Lu et al., 2020, Zhu et al., 2020), Africa has yet to fully employ sequence information to manage its COVID-19 outbreaks. This could be attributed to the limited competencies and infrastructural deficiencies (Jerving, 2020), such as low sequencing capacity, bioinformatics and computational skills and pipelines. To date, African countries have contributed few SARS-CoV-2 genomic data towards the global pool in growing open access repositories, such as NCBI GenBank and GISAID databases.

Molecular epidemiology studies remain imperative as African countries gradually reopen their borders with some level of COVID-19 testing but without real-time genomic surveillance to monitor the emergence of viral variants. To this end, this study pursued detailed phylogenetic inference, comparison of the evolutionary divergence, detection of repeat patterns and motifs, and analysis of the geographical distribution of SARS-CoV-2 trends in Africa. Here, we employed, among other methods, the Pango nomenclature system designed to implement a dynamic classification of SARS-CoV-2 lineages that incorporates both genetic and geographical components (O’Toole et al., in prep). The Pango nomenclature contains molecular signatures that are helpful for tracking SARS-CoV-2 introduction, emergence and spread (Andersen et al., 2020). We observed differences in the lineages circulating in Africa from those of most parts of the world. Lineage B.1.5 was identified as the dominant genetic lineage circulating in Africa, with most of the top 1% lineages in circulation worldwide not being found. This observation might be a consequence of under-surveillance as there is a relatively low number of African sequences available in the genetic databases, founder effects or the inefficient implementation of control measures, such as testing of travelers that may contribute to viral introduction and subsequent spread in the community. Consequently, the biological significance of the B.1.5 lineage, its epidemiologic features and spatial patterns deserve monitoring and further exploration, as there have been no reports of changes in transmissibility, fatality rates or vaccine efficacy, in contrast with lineage B.1.351 which dominated in early 2021 (Chen et al., 2021, Luo et al., 2021, Irfan and Chagla, 2021, Abu-Raddad et al., 2021, Shinde et al., 2021, Davies et al., 2021, Jassat et al., 2021) We also assessed the impact of the COVID-19 pandemic by evaluating the trends of absolute number of cases and deaths, but given the limited testing in Africa, we also relied on the proportion of the population that was screened for COVID-19 and the proportion that tested positive (positivity rate), which were relatively elevated for Mayotte, Guinea, South Sudan, and Tunisia. Compared to other continents, Africa appears to be relatively spared in terms of case fatality rate. Nonetheless, Egypt, Sudan, Chad, and Niger, all of which share borders, were found to have the highest numbers of COVID-19-related deaths, and thus further investigation is necessary to uncover the factors that led to this public health burden. We estimated that the impact of SARS-CoV-2 in Africa has been below the global average, both in terms of cases and mortality. However, this is based on the available information, that we found to be associated with the reported metrics and number of tests conducted, which are unfortunately very limited in Africa, as they are only conducted in those suspected to have interacted with a confirmed SARS-CoV-2 patient. Therefore, there is a high likelihood of underestimating cases, particularly by missing asymptomatic patients. This calls for policy decisions in Africa to be tailored towards expanding screening and improving implementation of measures that curb community spread. However, it is not certain that control measures that have proved effective in the global north will be equally effective in Africa. For instance, as shown in our findings, lockdown was not only counterproductive in different socio-economic areas, but also ineffective in curbing COVID-19 transmission in Africa. Some reasons that could be assigned include illiteracy, poverty, and cultural norms. Effective pursuance of grassroot education on good public health practices, mass distribution of disposable masks, free access to running water and soap, and availability of sanitizers in various public places, represent important avenues to tackle the current and future outbreaks.

We also identified de novo protein motifs that may have functional significance for SARS-CoV-2. Integrins are essential eukaryotic cells’ collagen receptors formed by a noncovalent interaction of two transmembrane glycoproteins subunits developing into about 24 variety of heterodimers, that facilitate the binding of cells to extracellular matrix and junctions, hence integrin-binding domain facilitates cell-attachment and cell-adhesion (Sigrist et al., 2020). Integrins may be used in place of the ACE-2 receptor because there is an integrin binding motif (arginine-glycine-aspartate [RGD]) on the spike protein (Beddingfield et al., 2021), that could potentially mediate viral entry into host cells and influence SARS-CoV-2 tissue tropism, viral transmission, and pathogenicity.

An N-glycosylation site employs a biosynthetic process of high complexity that is responsible for protein maturation along its secretory pathway (Yang et al., 2019, Galbán and Duckett, 2010). One of the major posttranslational modifications in proteins produced by eukaryotes, such as those on SARS-CoV-2, is glycosylation which determines protein configuration, function, and host adaptation. Glycosylations modify the three-dimensional structural orientation of proteins which is critical in protein-protein interactions of receptors and respective protein ligands. It also acts as a defense mechanism for SARS-CoV-2 against the immune cells and antibodies of the host, making it difficult to distinguish, identify and target the virus for elimination (Watanabe et al., 2019, Grant et al., 2020). Eleazar et al. (Ramírez Hernández et al., 2021) emphasizes how the level of glycosylation is a biologically significant mechanism affecting the post-translational modification of the viral surface proteins. Glycans play important roles in the viral life cycle, structure, immune evasion, and cell infection. This may in turn contribute to the cell infection rate and therefore to disease severity (Reily et al., 2019). We hypothesize that the presence of N-glycosylation domains on the SARS-CoV-2 genome may have implications for the binding affinity as previously described by Zhao et al. (Zhao et al., 2020).

The phylogenetic topological relationships revealed that African genomes tended to cluster with those from Europe, which is in line with the high cultural and business connectivity between these continents. We also observed several noteworthy outbreaks in Egypt, Democratic Republic of the Congo, Gambia, and South Africa, which reflects the higher number of sequences available for these countries. The higher viral diversity in Africa, compared to that in Oceania and South America, can initially be thought to be a reflection of the within-continent and inter-continent connectivity, as well as the travel patterns of individuals in Africa, which includes several major metropolitan areas, such as those in South Africa, Egypt, Ethiopia and Morocco. However, it might also be a consequence of diversity bottlenecks or the limited genomic surveillance in the continent. Despite being one of the few studies that comprehensively explored the viral genetic diversity, evolutionary history, and functional genome patterns of SARS-CoV-2 in Africa covering all regions in the continent, we understand that our observations might be conditioned by sampling bias. Additionally, our search for data brought to the fore a seeming lack of transparency in data disclosure and availability, both at the genetic and epidemiological fronts. This is concerning as it may deprive the continent and the global scientific community of useful information for consideration in the fight against the COVID-19 pandemic. The fact that the number of sequences generated from most of the countries were limited, and that some countries were not represented at all, might suggest that our observations may not represent the true state of the situation in the African continent, however, shedding light on the state of affairs may help inform public health policies. It is noteworthy that certain factors were believed to have contributed to the low or fewer number of sequences from Africa in public sequence databases. Key among these are the lack of resources and technical proficiencies. Sequencing, bioinformatics, and computational expertise can be greatly improved with capacity-building trainings organized by African entities and other international partners, such as the initiatives by the Fogarty International Center, National Institutes of Health and Johns Hopkins University Applied Physics Laboratory, that have been regularly training scientists from low and middle-income countries, particularly during the COVID-19 pandemic. These trainings have greatly improved the technical skills of participants towards analyzing the epidemiological and evolutionary trends of SARS-CoV-2 in Africa, as presented in this study. Furthermore, funding from African countries to support African scientists to carry out in-depth research on various aspects of SARS-CoV-2 in Africa is scarce (Oladipo et al., 2020). Besides the limited testing capacity in most African countries, full genome sequencing technology is also limited in most public health, research, or academic institutions. The associated cost, especially in less endowed countries, makes it an option not considered routinely. Local governmental commitment to funding research would allow scientists to be more independent in their research pursuits.

In conclusion, this work describes the molecular epidemiology, analyzes the genetic variability of SARS-CoV-2 in Africa, and highlights the need for continuous genomic and epidemiological surveillance, which is imperative for tracing the emergence of new genetic variants that can have significant effects on antigenicity, immunity, transmissibility, and potential vaccine escape. This information will also allow investigation of the transmission dynamics and resurgence of new waves of infection, as well as optimize public health measures, such as the deployment of vaccine formulations across the continent.

## Supporting information

Pangolin lineages circulating in Africa

GISAID Acknowledgement

Pairwise distance

## Data Availability

The data and script used for the analysis in this manuscript are available and can be access at:

https://github.com/Yinkaokoh/updatedSARCoV2_project

https://drive.google.com/file/d/1HMuhhR9xr7tWGcg8Tw0xkbkMKytB73Wa/view?usp=sharing

## Acknowledgments

We acknowledge the authors and laboratories responsible for generating and submitting sequences into GISAID’s EpiFlu Database. A full table of sequence authors is available in the supplemental information.

We also thank Helix Biogen Institute known as Helix Biogen Consult for their support. We gratefully acknowledge the Fogarty International Center at the National Institutes of Health and the Johns Hopkins University Applied Physics Laboratory for developing in-country capacity for whole genome sequencing and phylogenetics of SARS-CoV-2 and providing technical guidance. The content is solely the responsibility of the authors and does not represent official views of the National Institutes of Health.

## Data availability

https://github.com/Yinkaokoh/updatedSARCoV2_project

## SUPPLEMENTARY MATERIALS

### Supplementary Figures

**Supplementary Figure 1:**
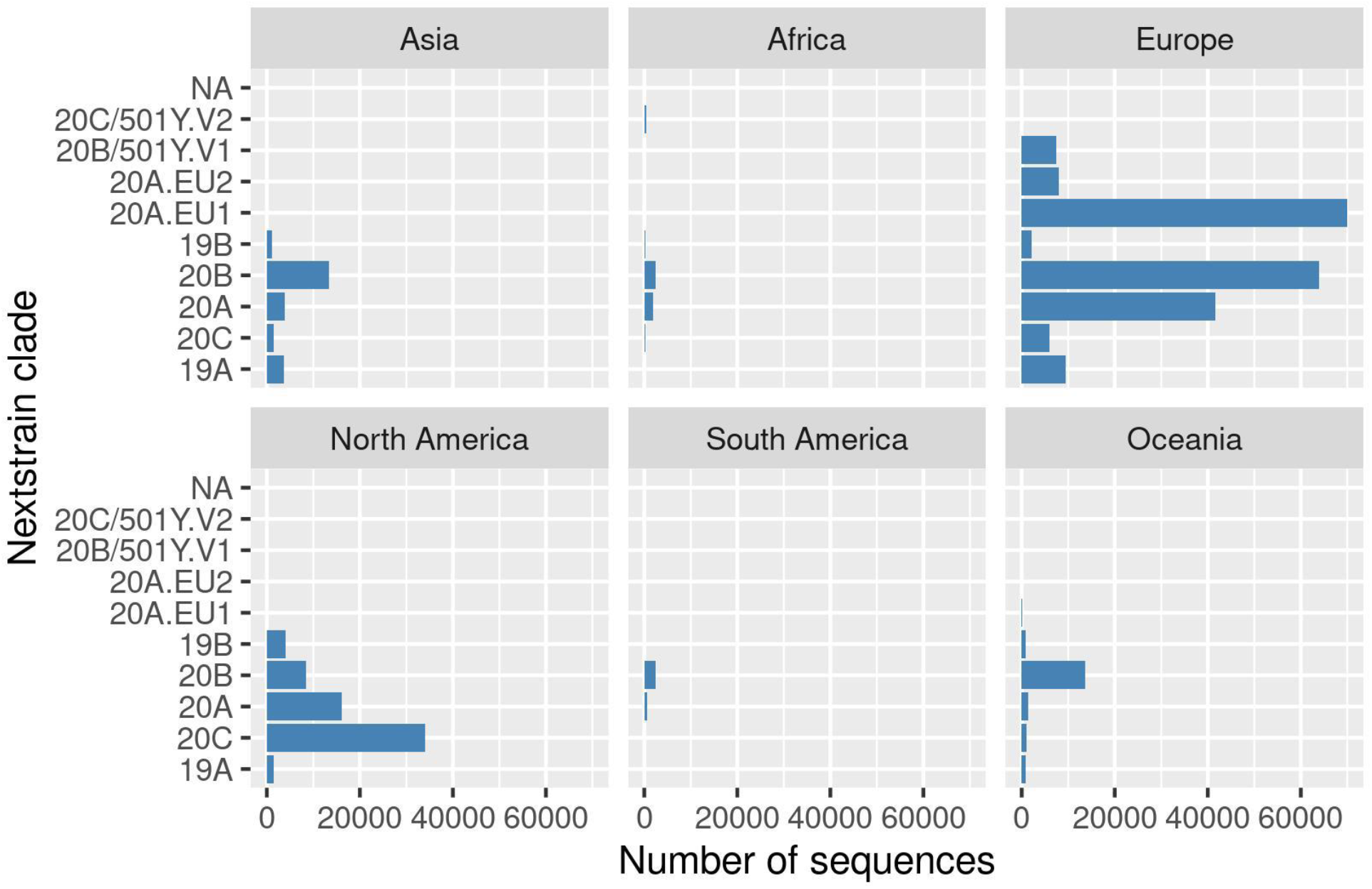
Diversity of Nextstrain clades across continents.

**Supplementary Figure 2:**
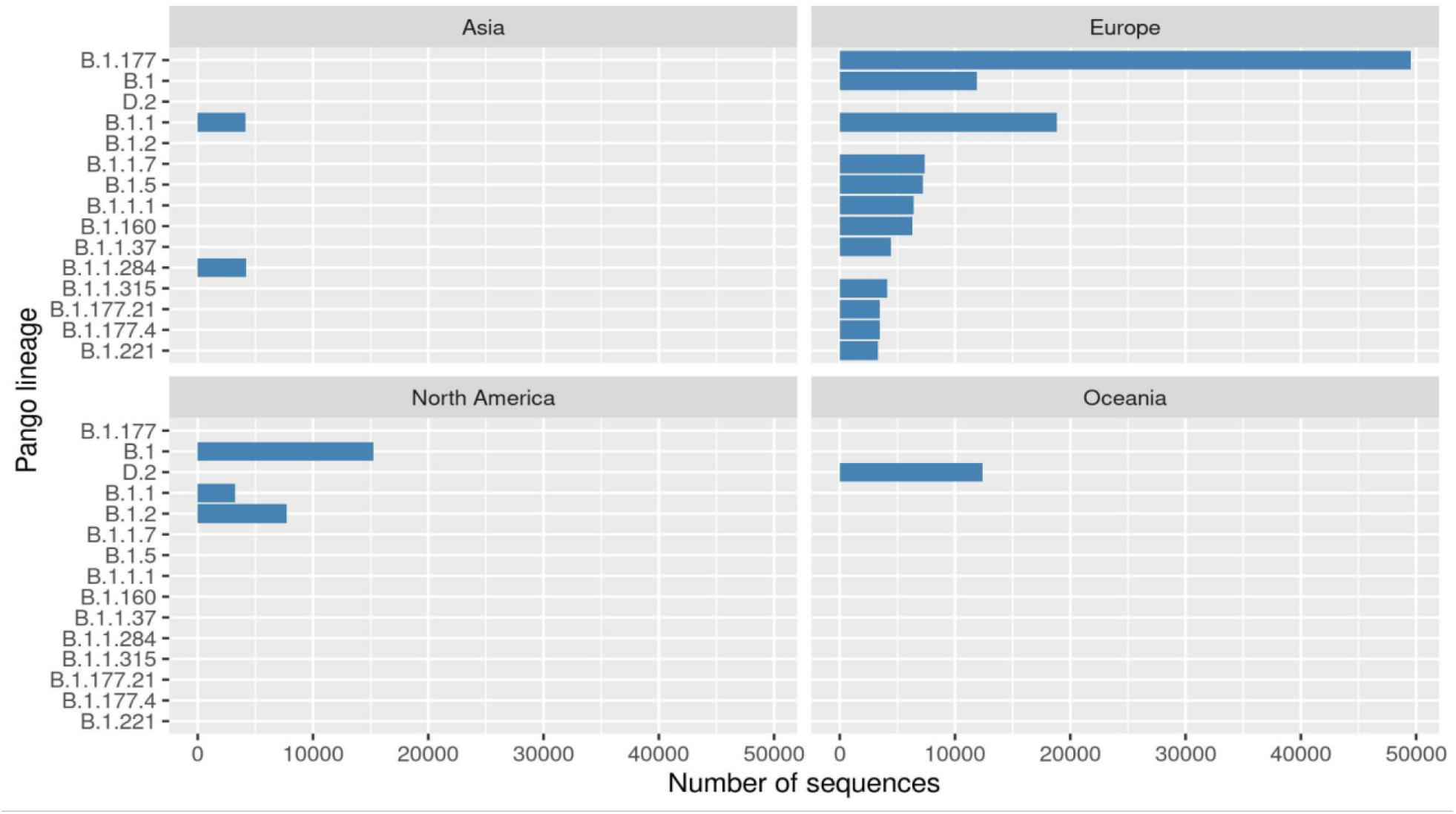
Top 1% Pango lineage diversity across continents.

**Supplementary Figure 3:**
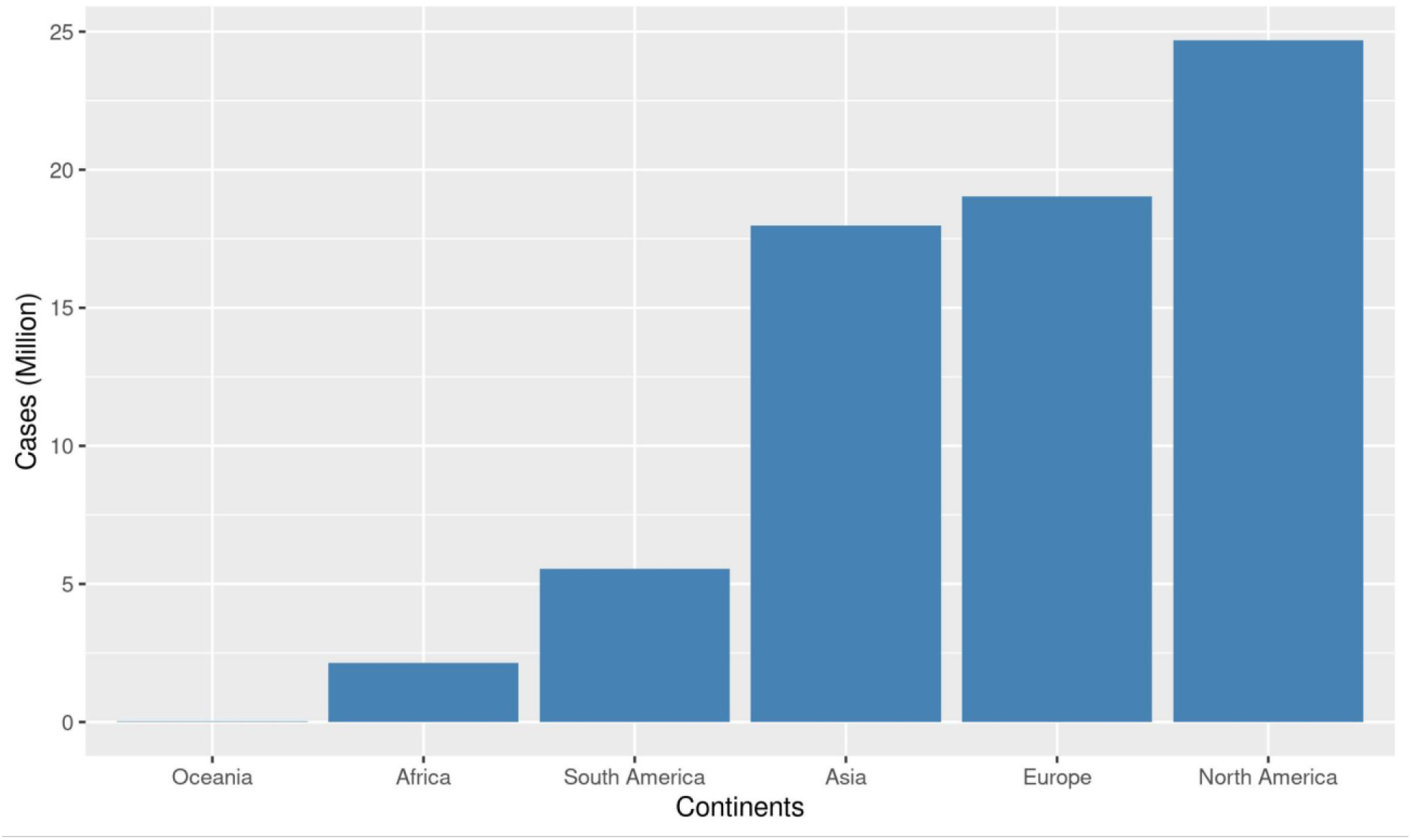
Absolute number of COVID-19 cases reported in the different continents.

**Supplementary Figure 4:**
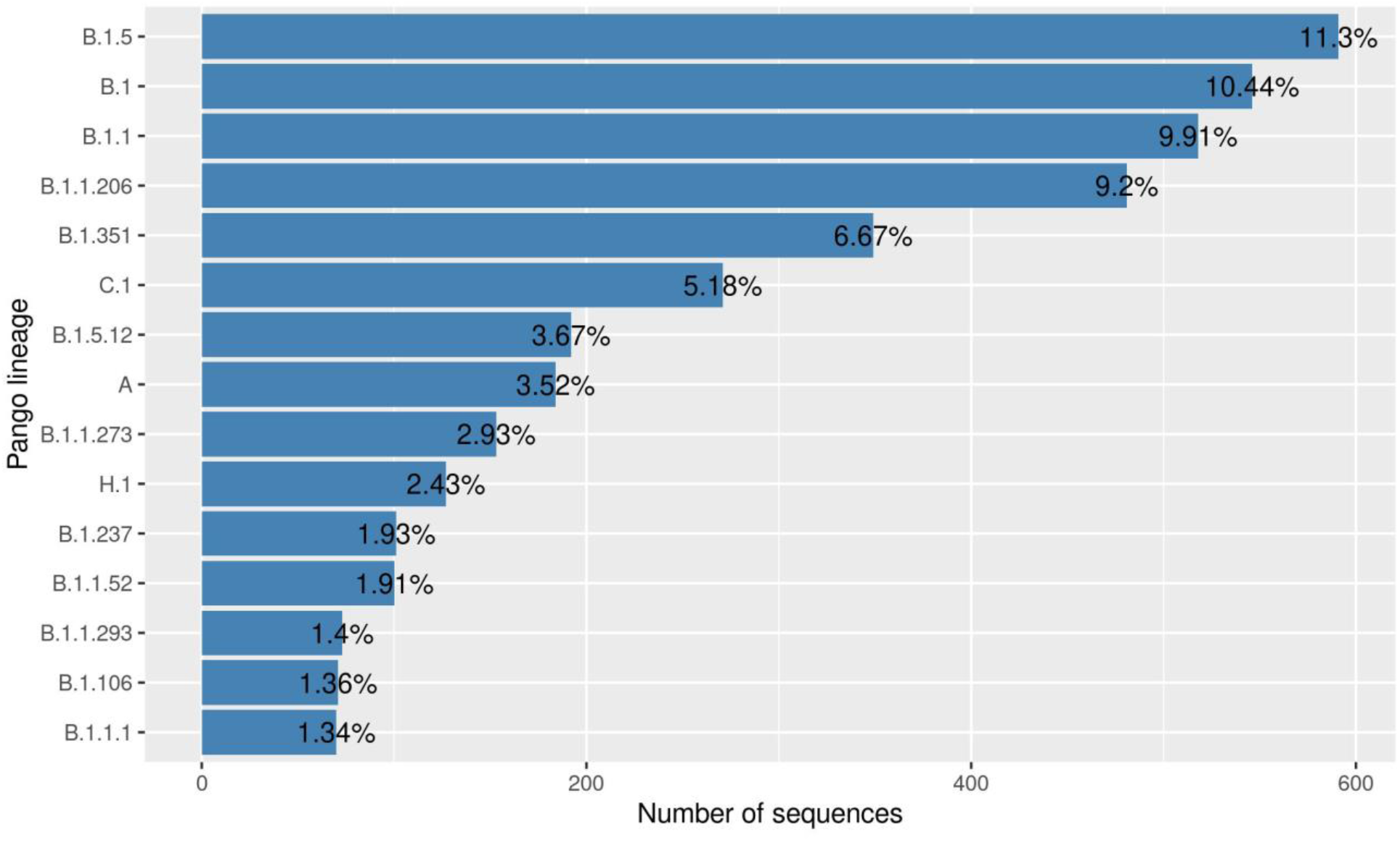
Top 10% Pango lineage diversity in Africa.

**Supplementary Figure 5:**
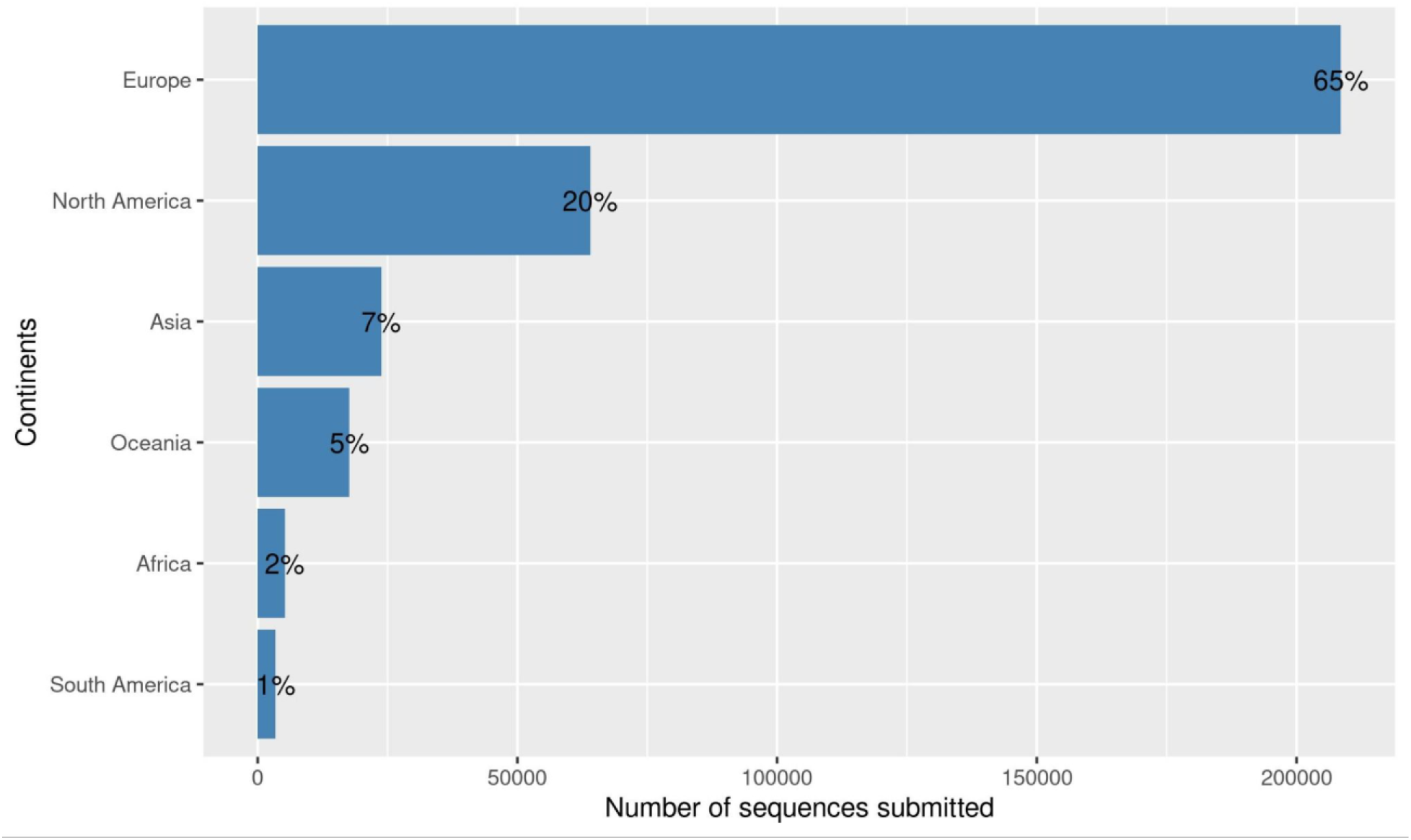
SARS-CoV-2 sequences submitted to GISAID. per continent (as of January 7, 2020).

**Supplementary Figure 6.**
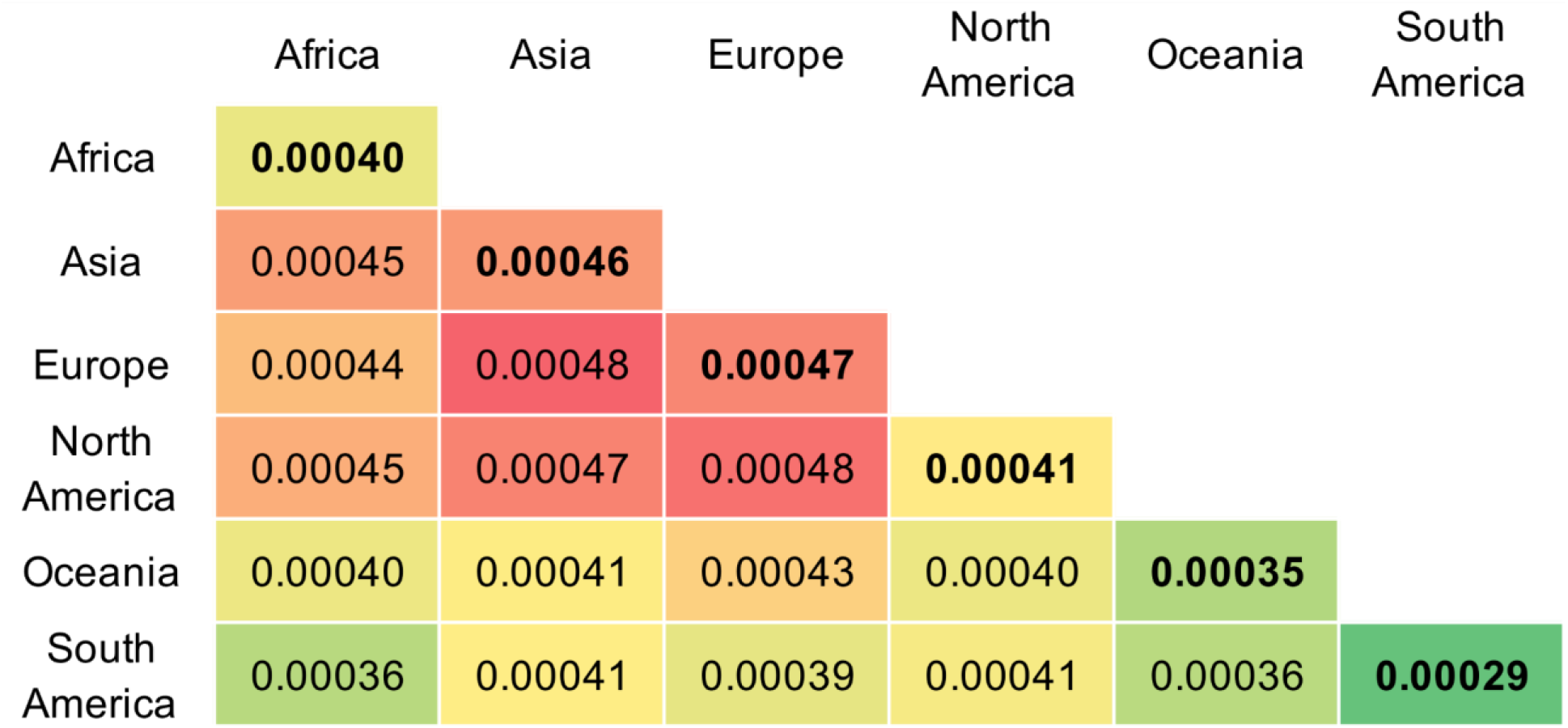
Evolutionary divergence of SARS-CoV-2 across continents. Pairwise matrix containing average pairwise genetic distances within (bolded in the diagonal) and between sequences from all continents. Green-to-red color scale highlights the most (red) and least (green) divergent within or between average distances.

Supplementary Figure 7. Maximum likelihood tree colored by continent. Phylogenetic tree inferred for a dataset with genetic sequences from all continents. Ultrafast bootstrap support is depicted by the circles at the nodes (smaller to larger width indicates low to high support).

### Supplementary Tables

**Supplementary Table 1: Pango lineages circulating in Africa**

**Supplementary Table 2: Accession numbers of sequences used in the study.**

**Supplementary Table 3:**
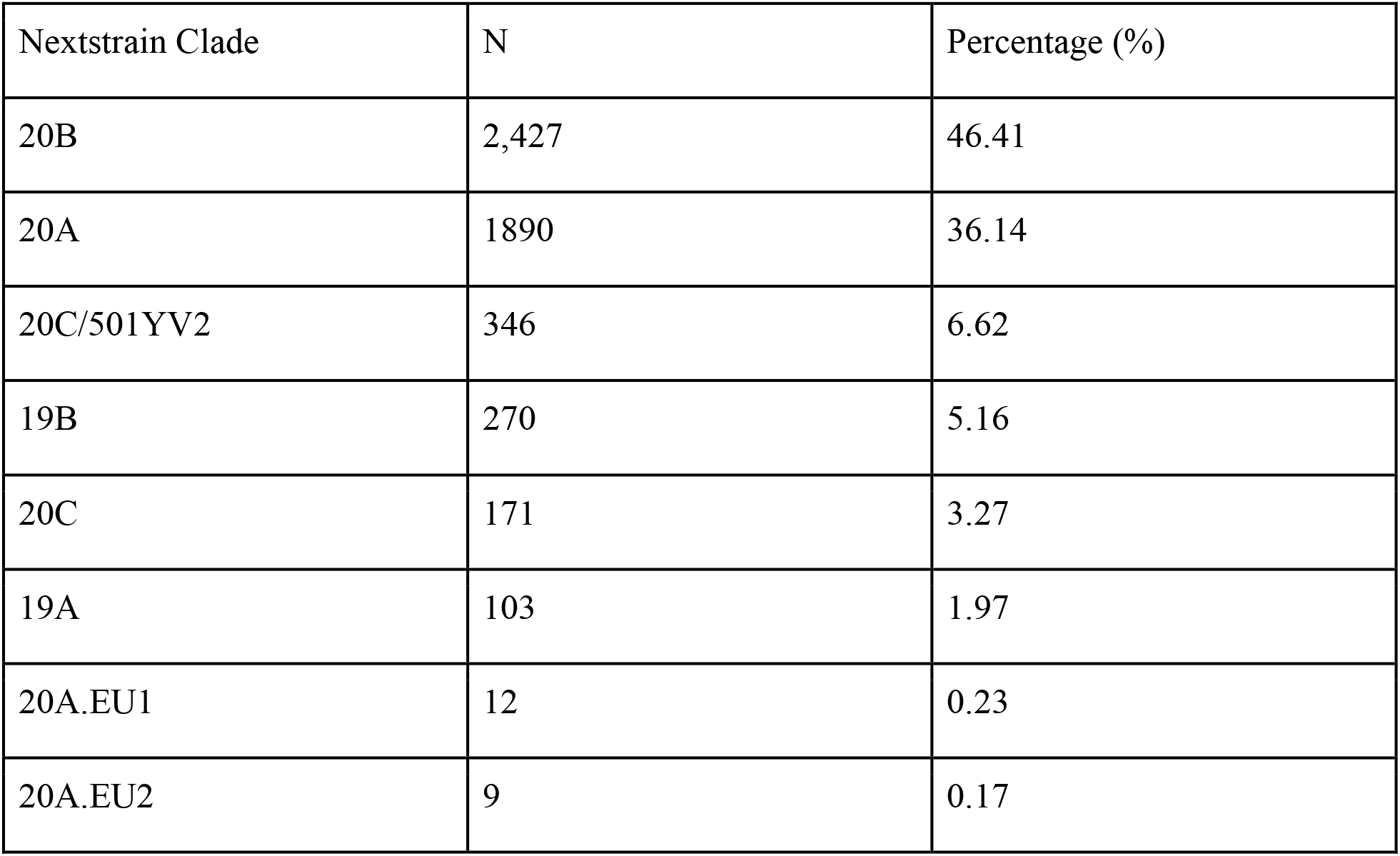
Nextstrain clades circulating in Africa.

**Supplementary Table 4: Pairwise genetic distance of SARS-CoV-2 isolates collected across continents**

